# A Cross-Modal Mutual Knowledge Distillation Framework for Alzheimer’s Disease Diagnosis: Addressing Incomplete Modalities

**DOI:** 10.1101/2023.08.24.23294574

**Authors:** Min Gu Kwak, Lingchao Mao, Zhiyang Zheng, Yi Su, Fleming Lure, Jing Li, Alzheimer’s Disease Neuroimaging Initiative

## Abstract

Early detection of Alzheimer’s Disease (AD) is crucial for timely interventions and optimizing treatment outcomes. Despite the promise of integrating multimodal neuroimages such as MRI and PET, handling datasets with incomplete modalities remains under-researched. This phenomenon, however, is common in real-world scenarios as not every patient has all modalities due to practical constraints such as cost, access, and safety concerns. We propose a deep learning framework employing cross-modal Mutual Knowledge Distillation (MKD) to model different sub-cohorts of patients based on their available modalities. In MKD, the multimodal model (e.g., MRI and PET) serves as a teacher, while the single-modality model (e.g., MRI only) is the student. Our MKD framework features three components: a Modality-Disentangling Teacher (MDT) model designed through information disentanglement, a student model that learns from classification errors and MDT’s knowledge, and the teacher model enhanced via distilling the student’s single-modal feature extraction capabilities. Moreover, we show the effectiveness of the proposed method through theoretical analysis and validate its performance with simulation studies. In addition, our method is demonstrated through a case study with Alzheimer’s Disease Neuroimaging Initiative (ADNI) datasets, underscoring the potential of artificial intelligence in addressing incomplete multimodal neuroimaging datasets and advancing early AD detection.

**Note to Practitioners:** This paper was motivated by the challenge of early AD diagnosis, particularly in scenarios when clinicians encounter varied availability of patient imaging data, such as MRI and PET scans, often constrained by cost or accessibility issues. We propose an incomplete multimodal learning framework that produces tailored models for patients with only MRI and patients with both MRI and PET. This approach improves the accuracy and effectiveness of early AD diagnosis, especially when imaging resources are limited, via bi-directional knowledge transfer. We introduced a teacher model that prioritizes extracting common information between different modalities, significantly enhancing the student model’s learning process. This paper includes theoretical analysis, simulation study, and realworld case study to illustrate the method’s promising potential in early AD detection. However, practitioners should be mindful of the complexities involved in model tuning. Future work will focus on improving model interpretability and expanding its application. This includes developing methods to discover the key brain regions for predictions, enhancing clinical trust, and extending the framework to incorporate a broader range of imaging modalities, demographic information, and clinical data. These advancements aim to provide a more comprehensive view of patient health and improve diagnostic accuracy across various neurodegenerative diseases.

## I. Introduction

A LZHEIMER’S disease (AD), a fatal neurodegenerative disorder, has become a pervasive global health challenge, with over 6.7 million Americans aged 65 and older affected in 2023 [1]. Despite decades of unsuccessful drug development, a new FDA-approved drug, Leqembi [2], and several drugs in testing with promising early results [3] offer a glimmer of hope. These pharmaceutical breakthroughs herald a new era in the fight against AD. Yet, the potential of these drugs to slow down disease progression is contingent upon early administration, ideally during the mild cognitive impairment (MCI) phase preceding AD dementia [4]. However, MCI is known to be heterogeneous, meaning that some individuals will convert to AD while others experience MCI due to non-AD-related brain diseases or conditions. Therefore, accurate differentiation of MCI patients who will convert to AD is crucial to ensure appropriate treatment at the optimal time [5].

Neuroimaging is a promising tool for early detection of AD [6]. Accurate detection of early-stage AD can benefit from the integration of multimodal neuroimaging datasets that capture brain structure and function from various perspectives, such as magnetic resonance imaging (MRI) and positron emission tomography (PET) [7]. However, such integration necessitates specialized dementia expertise, a valuable but scarce resource. Herein lies an opportunity for artificial intelligence (AI) to assist clinicians by integrating multimodal neuroimaging datasets to improve early AD detection capability.

However, a practical challenge in integrating multimodal neuroimages for early-stage AD detection is the disparate availability of different modalities among patients. While PET imaging provides highly valuable information for AD diagnosis, its availability is often limited due to several factors. For instance, while MRI is routinely used for AD-related clinical examinations in the U.S., PET may not be available for many patients due to its high cost, limited insurance coverage, and the need for specialized facilities and personnel [8]. Additionally, using radioactive tracers in PET scans introduces safety considerations that may further restrict its widespread use. Despite these limitations, PET provides valuable information about functional and pathological changes in the brain. For example, Florbetapir (FBP)-PET estimates beta-amyloid plaque density and is useful for early AD diagnosis [9]. The high predictive power of PET for early AD development is a key motivation for our work, as it allows us to leverage this valuable information even when working with patients who only have MRI data available.

There has been growing research interest in AI that addresses incomplete modalities in AD studies. This challenge is unique as the absence of a modality means losing all associated features. A common approach in the literature is to impute the missing modality, such as by leveraging autoencoders [10] or generative adversarial network (GAN) [11]. However, these methods were based on pre-extracted imaging features. We focus on image-based AI models using incomplete multimodal datasets to provide end-to-end predictions without prior feature engineering. Although several algorithms exist for integrating multimodal images [12], [13], limited works focused on tackling incomplete multimodal image datasets [14], [15].

To address this challenge, researchers have explored various approaches. These include not only traditional imputation methods but also more advanced approaches that leverage the complementary nature of different imaging modalities. Knowledge distillation (KD) has emerged as a promising method, gaining attention for its ability to transfer knowledge between models. In the context of multimodal learning, cross-modal KD has shown promise in transferring knowledge between different modalities [16]. Several AD studies have leveraged cross-modal KD to address the incomplete multimodal data challenge [14], [17], [18]. However, existing models focus on generating predictions using a single unified framework, regardless of whether patients have both PET and MRI or only MRI. This one-size-fits-all approach is not customized to each case.

To address this gap, we propose a novel framework that employs Mutual Knowledge Distillation (MKD) to effectively train models under incomplete modalities settings. MKD develops tailored predictive models for patient sub-cohorts with varying availability of image modalities (e.g., MRI-only or MRI&PET), optimizing prognostic capabilities across diverse patient populations. Different from existing KD methods that are primarily uni-directional, MKD involves a bi-directional knowledge exchange between the teacher and student models. The student learns from the higher predictive capacity of the teacher, which has access to multimodal imaging. Conversely, the teacher learns from the feature representation knowledge of the student, which has access to more samples for the more accessible modality. A key design for effective MKD is having a teacher model that can disentangle multimodal representations and use only the modality-common representation for classification. Figure 1 provides a high-level view of the proposed MKD framework applied to the context of AD diagnosis.

**Fig. 1:**
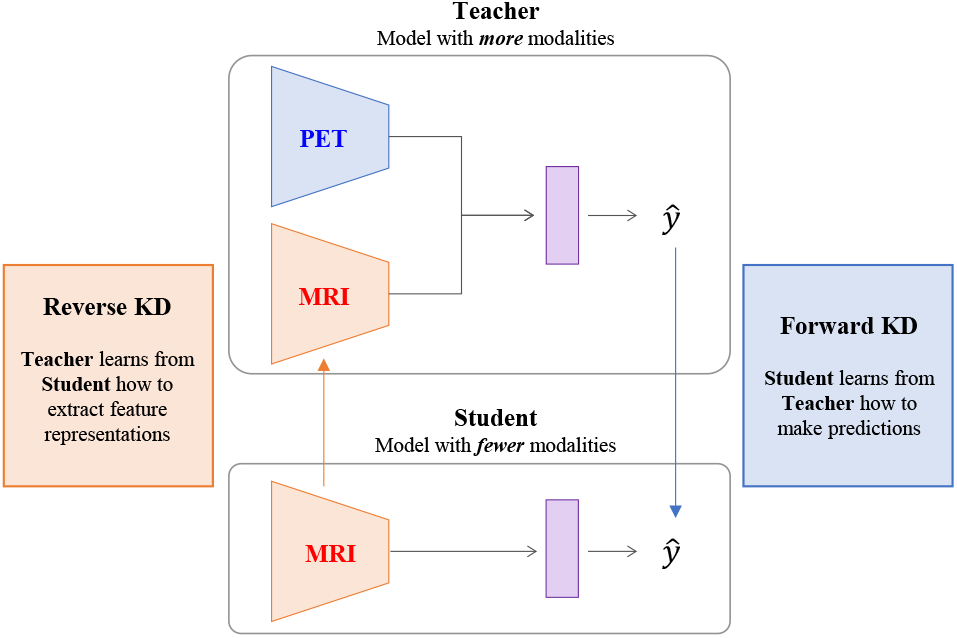
A conceptual depiction of the proposed MKD framework. The teacher, trained with more modalities, achieves higher accuracy, while the student, trained with fewer modalities but more samples, has better single-modal representation extraction capability. Both models enhance each other by mutually exchanging their respective strengths.

The main contributions of this paper are summarized as follows:

- We proposed a novel image-based AI framework using MKD to effectively train models under incomplete modalities settings. Different from existing methods that perform teacher-to-student KD [19], [20], our bi-directional MKD benefits both the teacher and the student. This framework develops distinct models for different patient groups based on their available imaging modalities, ensuring models are optimized for patients with either MRI-only or both MRI and PET data.
- We introduced a novel Modality-Disentangling Teacher (MDT) model within the MKD framework. The MDT employs multimodal information disentanglement to separate modality-common and -specific representations. It enables the teacher to capture robust predictive information shared by all imaging modalities and effectively transfer this knowledge to the student.
- We conducted a theoretical analysis to understand the effectiveness conditions of MDT. These theoretical findings, validated with simulation studies, demonstrate the benefit of multimodal information disentanglement for effective KD.
- We applied our MKD framework to the early detection of AD and achieved promising results compared to various state-of-the-art models. Our work showcases the potential of using AI to address the challenges of incomplete multimodal neuroimage datasets, paving the way for AI-empowered early AD detection.

## II. Related Works

### A. Multimodal Models

Multimodal neuroimages have been widely used in AD studies to develop AI models. Multimodal methods harness the complementary structural and functional information offered by diverse imaging modalities, resulting in higher predictive performances compared to single-modal methods [21]. For instance, Zhang et al. [22] proposed a multi-view method that employs low-rank tensor regularization to capture the high-order complementarity among MRI, PET, and genetic data. Another work introduced an efficient multimodal network with fewer parameters, employing a feature extractor that considers the relationship between structural MRI and its warp field characteristics followed by merging the extracted representations with clinical, demographic, and genetic data [23]. A recent multimodal fusion network employed discrete wavelet transformation to integrate structural and metabolic information, which were then used as inputs into classification models [24]. Recognizing that features extracted from different brain regions are inherently interrelated, an AD diagnostic model coupled interactions at both feature and modality levels to exploit the complementariness of multimodal neuroimaging data [25]. However, these studies do not take into account the challenge of incomplete multimodal data in AD diagnosis, where not every patient has access to all modalities.

### B. Cross-Modal Knowledge Distillation

Knowledge distillation (KD) is a technique for transferring knowledge from a complex teacher model to a simpler student model [26]. KD minimizes the Kullback-Leibler (KL) divergence between the teacher and student predictions, enabling a lightweight student to match a sophisticated teacher’s performance. In the context of AD, KD has been applied to enhance model accuracy in the face of constraints posed by small medical image datasets [27]. Additionally, an AD classification model was designed with structural distilling blocks incorporating multi-head self-attention for efficient data processing with MRI [28].

With the growing availability of multimodal data, KD has been applied to transfer knowledge across different modalities, forming a new area called cross-modal KD [16]. Typically, cross-modal KD aims to enhance the performance on a single target modality by leveraging other modalities. Cross-modal KD can be categorized into single-to-single and multi-to-single approaches. Figure 2 illustrates the differences between these KD approaches. Single-to-single cross-modal KD involves transferring knowledge from one modality to another. This approach has been applied in various domains, such as action recognition [29], brain tumor segmentation [30], and so on. Multi-to-single cross-modal KD has gained significant attention for its ability to leverage the high performance of multimodal teachers. This approach has been applied not only in computer vision [31] but also in medical domains. In brain tumor segmentation, Hu et al. [20] combined generalized KD with latent space constraints, enabling effective learning from a multimodal teacher when only the target modality is available. Several AD studies have leveraged cross-modal KD. A model predicting MCI conversion employed a multimodal multi-instance distillation scheme, addressing complex atrophy distributions via multi-instance probabilities [21]. A graph neural network was proposed to transfer knowledge from a graph-aware teacher to a graph-independent student, demonstrating improved AD progression prediction when graph information is unavailable [32]. However, most existing studies assume paired multimodal data, limiting their applicability in AD research and real-world clinical settings where incomplete multimodal datasets are common.

**Fig. 2:**
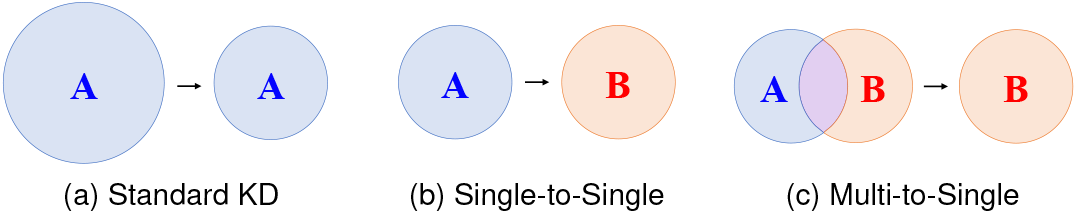
Categorization of KD methods: (a) Complex-to-simple standard KD within the same modality, (b) Single-to-single cross-modal KD, and (c) Multi-to-single cross-modal KD.

### C. Incomplete Multimodal Challenges

Recent studies have been introduced to tackle the challenge of incomplete multimodal data in AD studies. One approach is to impute the missing modality. An autoencoder-based model with graph regularization was proposed to impute the representations of the missing modality [10]. Several studies have utilized GANs for missing modality imputation. For instance, GAN was adopted to generate missing PET features based on the corresponding paired MRI features [11]. Another approach involved designing data-efficient loss functions that can navigate different missing patterns. Zhou et al. [33] leveraged complete multimodal samples to learn a common latent feature representation, while utilizing incomplete multimodal data from different subjects to learn modality-specific latent representations.

Several AD studies have leveraged cross-modal KD to address incomplete multimodal data challenges. Chen et al. [14] proposed a region-aware disentanglement module and an imputation-induced distillation module. Similarly, Wang et al. [17] presented a model utilizing KD to leverage supplementary information from all available data modalities. A joint learning framework integrating unsupervised cross-modal synthesis and AD diagnosis leveraged unpaired PET data to improve MRI-based diagnosis [18].

However, existing methods face several limitations. Most methods learn a single model, which generates predictions for all patients regardless of whether they have complete or incomplete modality data. This one-size-fits-all approach could be improved by having distinct models tailored to different modality availability scenarios. Additionally, cross-modal KD methods are primarily uni-directional, i.e., knowledge is transferred from the teacher to the student. We argue that there is also value in performing reverse KD, i.e., transferring knowledge from the student to the teacher, in incomplete multimodal settings.

### D. Summary of Gaps

The current landscape of AD research in incomplete cross-modal KD models highlights notable gaps. One primary concern is the prevalent use of a single prediction model for all patients, regardless of whether they have complete or incomplete modalities. We propose a framework that can efficiently learn models optimized for each diagnosis scenario (patients with only MRI vs. with MRI and PET) from incomplete multimodal data. Further, theoretical analysis on cross-modal KD remains limited. A recent theoretical analysis by Xue et al. [34] provided a key theoretical insight that the success of cross-modal KD hinges on the modality-common information, which inspired the design of our MDT model. However, this study was restricted to single-to-single cross-modal KD. In our work, We extended this theoretical framework to the multi-to-single setting. Overall, this paper introduces novel strategies for modeling with incomplete modalities and enhancing the cross-modal KD process for AD diagnosis.

## III. Proposed Method

Our MKD framework will be presented in the context of two imaging modalities: MRI and PET. MRI is commonly used in AD assessments, but PET scans are less frequent due to higher cost [8]. This creates two patient sub-cohorts: those with only MRI and those with both MRI and PET. Our objective is to develop accurate models for each sub-cohort. Specifically, the teacher model is designed to predict using both MRI and PET, while the student model utilizes only MRI data.

The MKD framework involves three sequential training steps. First, we develop a teacher model that extracts modality-common representation, namely the Modality-Disentangling Teacher (MDT). Second, we train a student model, termed MDT-Student, through conventional classification error minimization and “forward KD” from the MDT. Lastly, we train a new teacher, MDT^+^, by leveraging the student’s MRI feature extraction capabilities. We call this process “reverse KD” as knowledge flows from the student to the teacher. An overview of this process is shown in Figure 3.

**Fig. 3:**
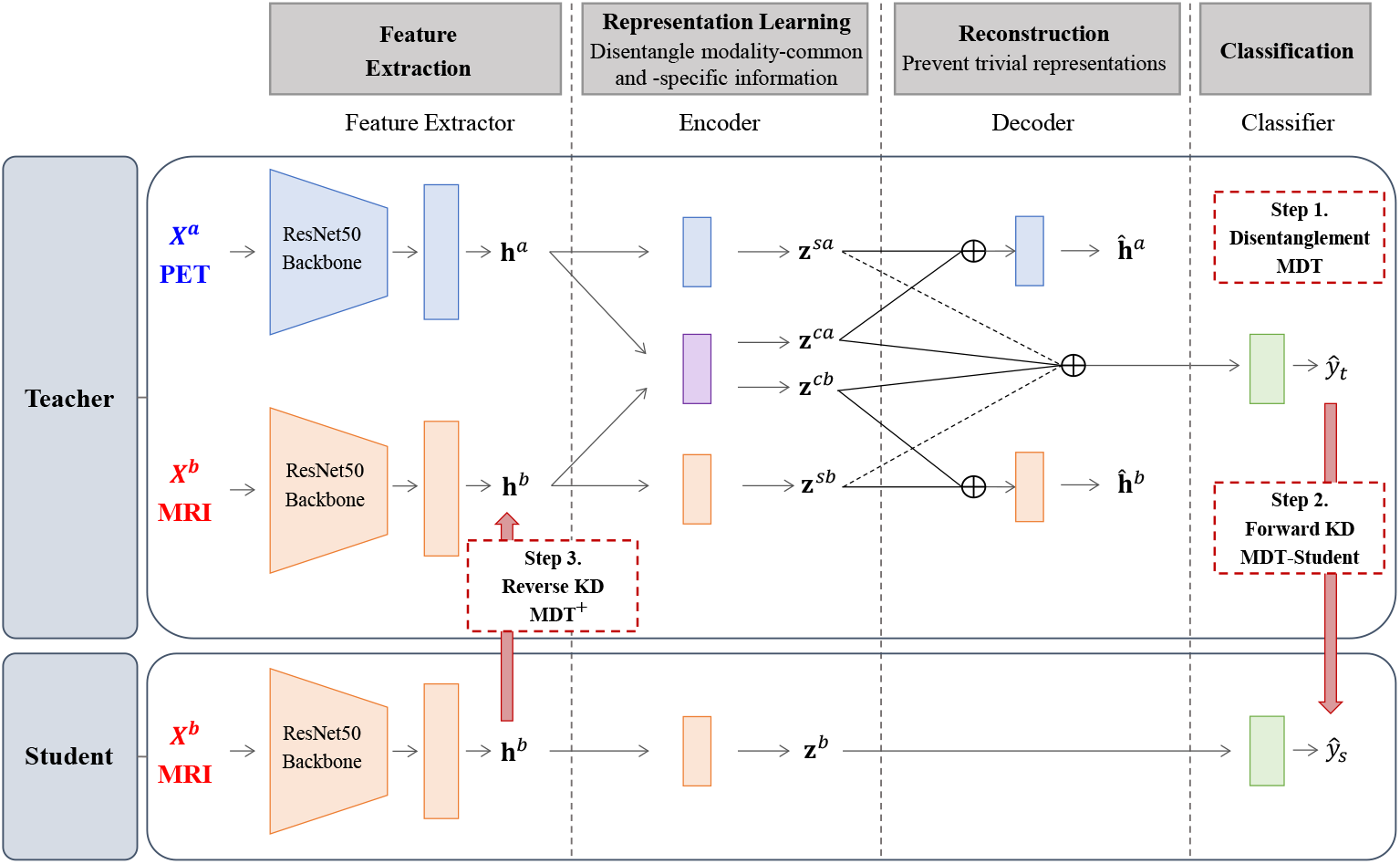
Teacher and student model architectures and training process in the MKD framework. MDT/MDT^+^: upper architecture without/with dash lines; MDT-Student: bottom architecture. First, the teacher is designed to disentangle modality-common and -specific information and uses modality-common information for classification (MDT). Second, the student then learns from the teacher through forward KD (MDT-Student). Last, the teacher is subsequently updated by learning the student’s feature extraction capability through reverse KD, and adding modality-specific information (MDT^+^).

In what follows, we describe the three training steps in Sections III-A, III-B, and III-C. Section III-D presents a theoretical analysis on multi-to-single cross-modal KD.

### A. Modality-Disentangling Teacher (MDT)

Contrary to existing methods that focus on training a highly accurate teacher for KD, recent insights suggest that the efficiency of cross-modal KD hinges on the strategic preservation and transfer of essential, modality-common information from the teacher to the student [34]. Thus, we begin by developing a teacher model that learns to disentangle modality-common and modality-specific representations from multimodal datasets. This disentanglement process not only simplifies the classification task by reducing dimensionality and removing redundant or noisy information from the input modalities, but also allows extracting the modality-common representation that is essential for an effective forward KD. Next, we introduce the details of the MDT model.

Consider the case of two input modalities such as PET and MRI images, denoted as **X**^*a*^ and **X**^*b*^. The task is a binary classification of an MCI patient into an AD converter or a non-converter (*y* = 1 or 0). The architecture of the MDT encompasses several subnetworks, including feature extractors, encoders, decoders, and a classifier.

To begin, a feature extractor maps the image of a modality **x**^*m*^ to a latent vector **h**^*m*^, *m* ∈ {*a, b*}. This subnetwork can adopt any architecture capable of handling imaging data as the backbone, such as ResNet-50 [35], followed by a global average pooling layer and a fully connected projector layer.

Following feature extraction, each latent vector **h**^*m*^ passes through two encoders in a parallel manner. The first is the modality-specific encoder that transforms the latent vector of a given modality **h**^*m*^ into a modality-specific representation, denoted as **z**^*sm*^, capturing information unique to each modality. The second encoder is a modality-common encoder that takes as input the latent vector of both modalities (**h**^*a*^, **h**^*b*^) and outputs the modality-common representation, **z**^*cm*^, extracting shared information across both MRI and PET images. These encoders are implemented as simple feed-forward neural network layers. To ensure consistency in representation scales across different modalities, all the representations undergo L2 normalization.

The obtained **z** representations are then combined for reconstruction and classification tasks. For reconstruction, the sum of modality-common (**z**^*cm*^) and modality-specific (**z**^*sm*^) representations of each modality is fed into the corresponding decoder to reconstruct **h**^*m*^, denoted as **ĥ**^*m*^. These reconstruction tasks prevent extracting trivial representations. For classification, only the sum of modality-common representations (**z**^*cm*^) is passed to a classifier to predict the label *y*. As shown in our theoretical analysis and experimental results, using only modality-common representations for classification facilitates effective teacher-to-student KD.

To jointly learn disentangled representations and classification, the MDT utilizes a combination of losses, including similarity, difference, reconstruction, and classification losses:

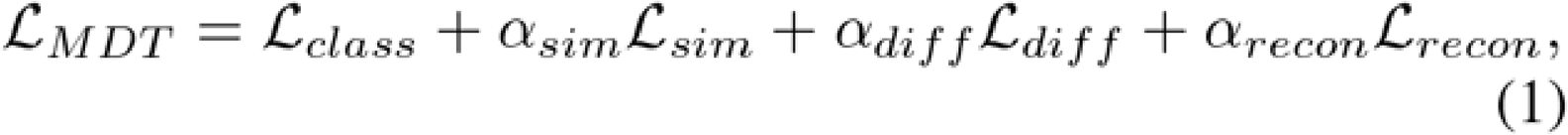

where each *α* denotes a balancing hyperparameter.

ℒ_*sim*_ aims to maximize the similarity between modality-common representations extracted from each modality. This helps align representations from different modalities in a shared subspace:

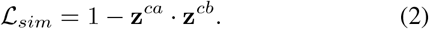

Herein, the inner dot product is equivalent to the cosine similarity as **z**^*ca*^ and **z**^*cb*^ are L2-normalized. We added one to the negative cosine similarity to make the minimum value of ℒ_*sim*_ as zero for simplicity.

ℒ_*diff*_ ensures that modality-common and -specific representations capture distinct aspects of the input. Redundancy is avoided by enforcing a soft orthogonality constraint between modality-common and -specific representations within each modality as well as between the modality-specific representations across different modalities:

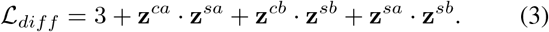

By adding three to the sum of inner products, ℒ_*diff*_ is adjusted to ensure non-negativity.

ℒ_*recon*_ helps ensure encoders extract representative information from each modality instead of trivial features. Reconstruction error is measured as the mean squared error (MSE) between **h**^*m*^ and **ĥ**^*m*^:

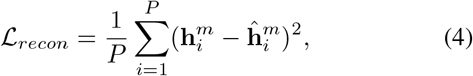

where *P* is the dimension of **h**^*m*^ and **ĥ**^*m*^.

Lastly, we employ cross-entropy (CE) loss for classification:

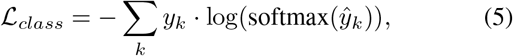

where *ŷ*_*k*_ is the predicted logit of the *k*-th class and *k* ∈ {0, 1}.

### B. Forward KD: Student Learns from MDT (MDT-Student)

In this section, we discuss the student model architecture and the forward KD process. Recall that the teacher uses two modalities *X*^*a*^ and *X*^*b*^ (PET and MRI) whereas the student focuses on predicting MCI conversion to AD using only *X*^*b*^ (MRI). To facilitate KD between models, the student architecture is designed to resemble the branch of the teacher that involves *X*^*b*^. Specifically, the student includes a feature extractor, an encoder, and a classifier. There are three remarks we want to highlight: First, the student is equipped with only one encoder and does not conduct disentanglement between modality-common and -specific representations because it only receives *X*^*b*^ as input. Second, the student architecture does not include a decoder for reconstruction. While adding a decoder is straightforward, we found in our experiments that a decoder did not improve the classification performance of the student, which may be due to the trade-off between reconstruction and classification. Thus, we decided to not include a decoder for simplicity. Third, the student’s feature extractor uses pre-trained weights from the teacher for utilizing its multimodal classification strengths. To utilize pre-trained weights from the teacher model, it is imperative that the representations input into the student’s classifier are at similar scale. The teacher model ensures consistent scales by L2-normalizing **z**^*ca*^ and **z**^*cb*^. To preserve consistent scales in the student model, we doubled the output of the student encoder before inputting it into the student classifier.

MDT-Student is trained by minimizing the following loss:

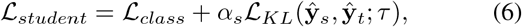

where ℒ_*KL*_ is the KL divergence loss that measures the difference between predicted logits of the student and teacher, **ŷ**_*s*_ and **ŷ**_*t*_, respectively. *τ* is the scaling hyperparameter for logits and it allows the student to learn the dark knowledge of the teacher [26]. *α*_*s*_ is a hyperparameter of balancing the KL divergence loss for training the student.

### C. Reverse KD: Teacher Learns from MDT-Student (MDT^+^)

The last step is to obtain a new teacher model (MDT^+^) by learning from the MDT-Student. In incomplete multimodal settings, the student model, being exclusively trained on a single modality **X**^*b*^, can utilize more samples than the teacher. While the inability to use **X**^*a*^ might compromise its classification performance, the student model excels at extracting feature representations from **X**^*b*^. This capability can be exploited to develop an improved teacher model, which we call MDT^+^.

The training procedure of MDT^+^ is similar to MDT but with three key differences. First, the weights of the feature extractor of this new teacher are initialized using the weights from the corresponding subnetwork of MDT-Student, while the weights for the remaining subnetworks are randomly initialized. Second, an additional KD loss to enable the transfer of the feature extraction knowledge of **X**^*b*^ from the student to the teacher. We employed the following loss function for performing representation-level KD:

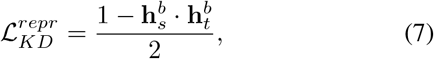

where 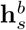 and 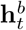 denote the outputs of modality *b* from the feature extractors of the student and teacher. 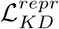 maximizes the cosine similarity between 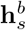 and 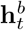 [36], and is added to (1) with a balancing hyperparameter 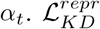 is divided by two to scale the loss from zero to one, ensuring training stability. Further, the classifier of MDT^+^ uses both modality-common and modality-specific representations for predicting the label to optimize predictive performance.

At test time, the trained MDT-Student will be used to predict for patients with only MRI and MDT^+^ will be used to predict for patients with both MRI and PET.

Algorithm 1 provides pseudo code of training the models in Sections III-A, III-A, and III-C in the proposed MKD framework. Note that we also explored a joint training approach. However, it led to poor disentanglement of modality information and worse student model performance. Based on our results, we found that the sequential training approach allows each step to focus on its specific objective, effectively leveraging multi- and single-modal data strengths.

The balancing hyperparameters, *α*_*sim*_, *α*_*diff*_, *α*_*recon*_, and *α*_*t*_ can be tuned via a grid search. Since the most critical component of the MDT is representation disentanglement, we suggest focusing on tuning hyperparameters that directly influence the ability to disentangle, *α*_*sim*_ and *α*_*diff*_. From our experiments, we did not find *α*_*recon*_, *α*_*s*_, and *α*_*t*_ critically affect model performance. Thus, we adjusted them so that the scale of respective losses would be aligned with the CE loss. Hyperparameter tuning details can be found in Appendix C.

### D. Theoretical Analysis

In this section, we present our theoretical analysis that shows a multimodal teacher that emphasizes the modality-common representations provides more effective knowledge transfer compared to a regular teacher that uses both modality-common and modality-specific representations for prediction. The theoretical analysis is presented in two parts: (1) the effectiveness of the MDT teacher-to-student KD is positively associated with the proportion of modality-common features over the combination of modality-common and modality-specific features, and (2) the student trained using MDT exhibits a lower upper bound on the empirical KD loss compared to the student trained with a regular teacher.

To begin, we describe the concept of a modality Venn diagram [34] to help understand the dynamics of how multimodal data is generated. Let 𝒳^*a*^, 𝒳^*b*^, and 𝒴 be the feature space of modality *a*, modality *b*, and the label space, respectively. Consider a sample (**x**^*a*^, **x**^*b*^, *y*) drawn from an unknown distribution P that spans 𝒳^*a*^ × 𝒳^*b*^ × 𝒴. According to the modality Venn diagram, a sample (**x**^*a*^, **x**^*b*^, *y*) is generated from a latent subspace represented as (**z**^*c*^, **z**^*sa*^, **z**^*sb*^, *y*) ∈ 𝒵^*c*^ × 𝒵^*sa*^ × 𝒵^*sb*^ × 𝒴, where *c, sa*, and *sb* denote modality-common, modality a specific, and modality b-specific latent features, respectively. The generation process is based on the following rules:

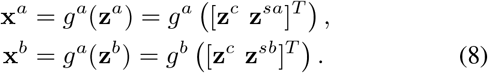

#### Algorithm 1 Mutual Knowledge Distillation (MKD) Framework

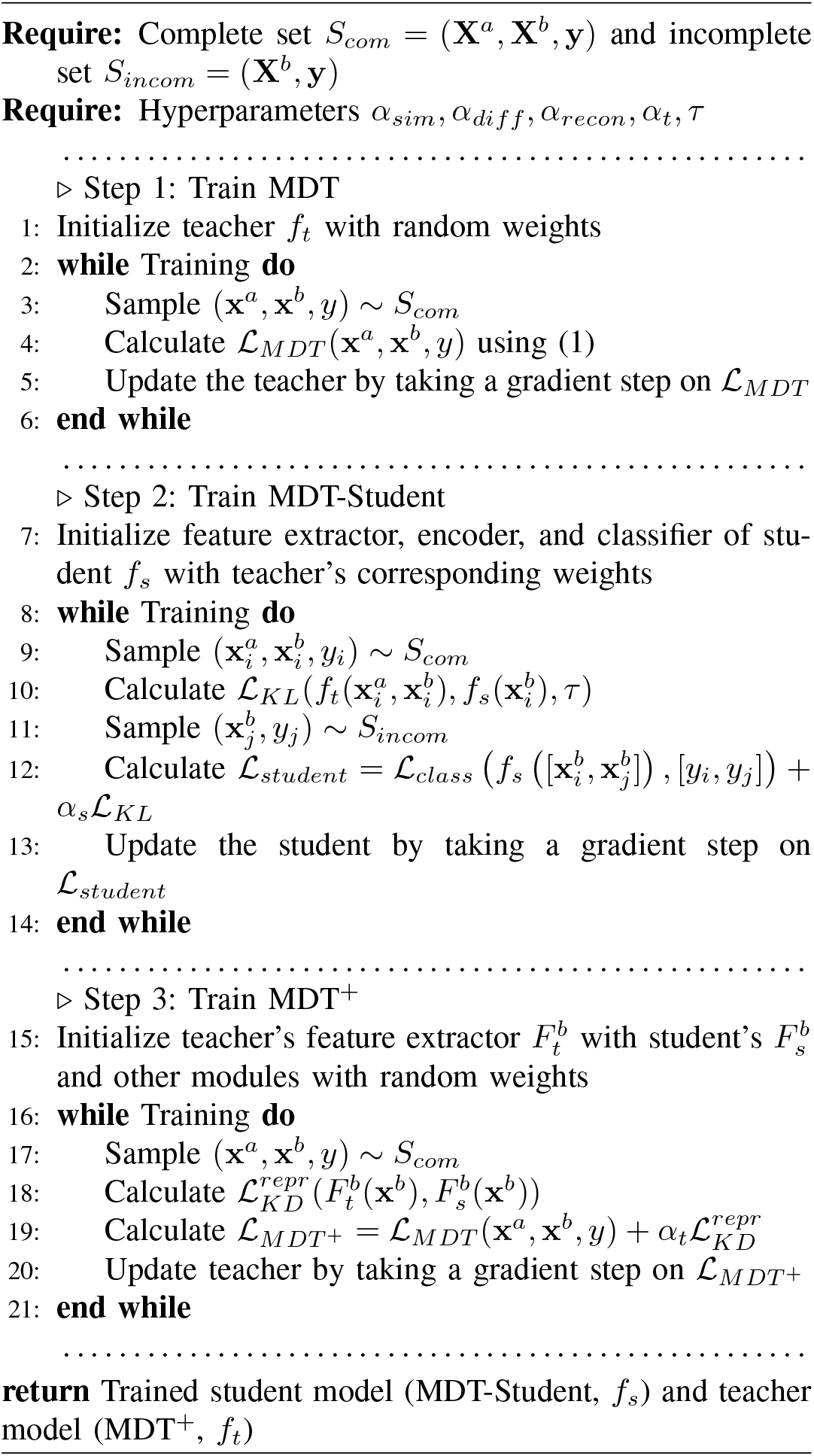

Herein, for 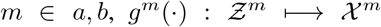 denotes an unknown generating function. For simplicity, we assume that the latent subspace linearly determines the label *y*.

Let 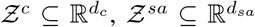, and 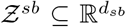. Define the union of the latent subspace as 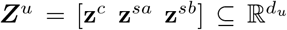, so that *d*_*u*_ = *d*_*c*_ + *d*_*sb*_ + *d*_*sb*_. Denote the ratios of modality common, modality *a*-specific, and modality *b*-specific features over all features as *γ, α*, and *β*, respectively (i.e., *γ* = *d*_*c*_*/d*_*u*_, *α* = *d*_*sa*_*/d*_*u*_, *β* = *d*_*sb*_*/d*_*u*_), with *γ* + *α* + *β* = 1 and *γ, α, β* ∈ [0, 1]. We provide the following Theorem 1 to bound the empirical KD loss of MDT-Student in terms of *γ*. This theoretical result shows that with a given *α*, the upper bound of the empirical KD loss decreases as *γ* increases. See Appendix A for detailed proof.

#### Theorem 1

(Bound on the MDT-Student Empirical KD Loss): Consider a linear teacher model that predicts using modality-common representation, i.e.,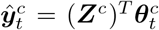; and a linear student model, i.e., 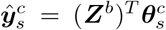 The KD loss is defined by the KL divergence between the predictions from the student and teacher:

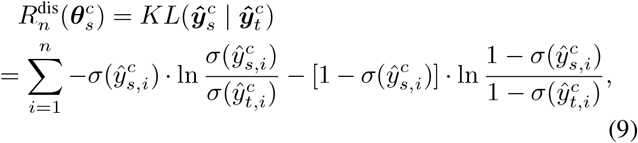

where *σ*(·) is the sigmoid function. Suppose max ***Z***^*u*^(***Z***^*u*^)^*T*^, (***Z***^*u*^(***Z***^*u*^)^*T*^)^−1^ ≤ *λ*, holds for *m* ∈ {*a, b, c*}. If there exists (*ε, δ*) such that 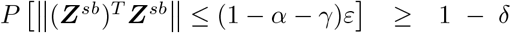, then with probability at least 1 − *δ*, the empirical KD loss is upper bounded by

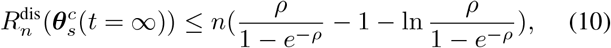

where *ρ* = *λ*^1.5^(*λ*^2^ + 1)(1 − *α* − *γ*)*ε*.

Next, we show that the student trained with MDT is expected to have a more effective KD than the student trained with a regular teacher by showing that the former has a tighter upper bound on empirical KD loss. See Appendix B for proof.

#### Theorem 2

(Comparing the empirical KD losses between MDT and a regular teacher): Under the same setting as Theorem 1. Additionally, suppose max ***Z***^*u*^(***Z***^*u*^)^*T*^, (***Z***^*u*^(***Z***^*u*^)^*T*^)^−1^ ≤ *λ*, where *u* denotes the union set of all the feature representations. Consider two teachers: MDT that predicts using only modality-common representations, i.e.,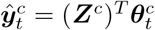 ; and a regular teacher which predicts using both modality-common and modality-specific representations, i.e.,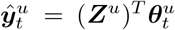. Denote ℳ^*c*^ and ℳ^*u*^ as the upper bound on empirical KD loss of the student with MDT and the regular teacher, respectively. Then, with probability at least 1 − *δ*, ℳ^*u*^ ≥ ℳ^*c*^.

## IV. Simulation Experiments

We conducted simulation experiments to evaluate the MKD under multimodal settings with varying amounts of modality-common information and the levels of incomplete ratios. A unique trait of the proposed model is the bi-directional KD design that allows two models to learn from each other’s strengths. Thus, we assess the performance of both the student and the teacher models compared to their baselines. To validate the importance of modality-common information in the effectiveness of KD, which is foundational to the design of MDT, we compare MDT with a regular multimodal teacher that does not emphasize modality-common information.

### A. Simulation Data Generation

We consider two sub-cohorts of virtual patients: one with two modalities available (**X**^*a*^, **X**^*b*^) and the other has one modality **X**^*b*^. For each sample (a virtual patient), we first assigned the label *y* of the sample (1 for converter and 0 for non-converter). Upon establishing the patient’s condition, we proceeded to generate latent representations **z**^*c*^,**z**^*sa*^, and *z*^*sb*^, where 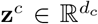 is the modality-common representation and 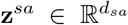 and 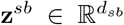 are the modality-specific representations for **X**^*a*^ and **X**^*b*^. These representations were independently sampled from normal distributions conditioned on the patient’s status *y*, ensuring disentanglement of the representations, a core assumption of our model. Specifically, the latent representations were generated by **z**^*m*^ ∼ *𝒩* (*µ*_*y*_, Σ_*m*_) where *m* ∈ {*c, sa, sb*} and *µ*_*y*_ and Σ_*m*_ denote the mean vector and the covariance matrix. We set *µ*_0_ = 0.0 and *µ*_1_ = 0.5. We constructed Σ_*m*_ with its elements defined as Σ_*m*_[*p, q*] = *ρ*^|*p*−*q*|^*σ*^2^ for *p, q* = 1, …, *d*_*m*_. We set *ρ* = 0.5 and *σ*^2^ = 1.0, inducing feature correlations. The proportion of modality-common representation is computed as *γ* = *d*_*c*_*/*(*d*_*c*_ + *d*_*sa*_ + *d*_*sb*_). We simulated multimodal settings with varying proportion of modality-common information, *γ*, by altering *d*_*c*_ in values of 2, 6, …, 22, while maintaining *d*_*c*_ + *d*_*sa*_ + *d*_*sb*_ = 30. This modification resulted in *γ* values ranging from 0.07 (i.e., modalities are highly unrelated) to 0.73 (i.e., modalities are highly overlapping).

Then, we transformed the representation vectors into observed data through matrix multiplication with a non-linear activation function. Specifically, the transformation applied was **x**^*m*^ = LeakyReLU(**W**^*m*^ · **z**^*m*^), where 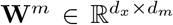 is a randomly initialized transformation matrix. *d*_*x*_ is the dimension of the observed data vector and was equally set to 20 for **x**^*sa*^, **x**^*sb*^, and **x**^*c*^. The slope of LeakyReLU was set to 0.5. The final samples for each modality, **X**^*a*^ and **X**^*b*^, were constructed by concatenating the modality-common vector **x**^*c*^ with modality-specific vector **x**^*sa*^ and **x**^*sb*^, respectively. Namely, the final data matrix **X**^*a*^ was formed by concatenation as **x**^*c*^ ⊕ **x**^*sa*^ and for **X**^*b*^ as **x**^*c*^ ⊕ **x**^*sb*^. Finally, utilizing the process, we generated a complete modality dataset (**X**^*a*^, **X**^*b*^, **y**) with *N*_*com*_ samples and an incomplete modality dataset (**X**^*b*^, **y**) with *N*_*incom*_ samples. Both datasets were created ensuring a 1:1 class distribution.

### B. Simulation Model Implementation Details

The architecture of the MDT begins with a ten-dimensional single-layer feature extractor. The encoders encompassed a five-dimensional layer, Layer Normalization, a ReLU activation, and another five-dimensional layer. The decoders mirrored the encoders. A single-layer classifier was employed.

Regarding the training hyperparameters, the batch size was fixed to 16 for all models. AdamW optimizer [37] with a weight decay of 0.0001 and half-cosine learning rate scheduling was employed. The teacher was trained for 100 epochs with a learning rate of 0.001, while the student was trained for 30 epochs with a learning rate of 0.0001. We set *α*_*sim*_ = 2.0, *α*_*diff*_ = 1.0, *α*_*recon*_ = 100.0, *α*_*s*_ = 100.0, and *α*_*t*_ = 5.0 after hyperparameter tuning. A grid search was conducted over *τ* ∈ {0.1, 0.5, 1.0, 5.0, 7.0, 10.0}, commonly used in KD studies [26]. The value of *τ* was set to 5.0, which yielded the best MDT-Student performance. In our method’s hyperparameter tuning process, the focus is primarily on representation disentanglement. To address this, we outlined an efficient process for tuning multiple balancing hyperparameters in Appendix C.

### C. Student Performance Evaluation

This experiment aims to assess the utility of the MDT design, which emphasizes modality-common information, to improve the student. Three student models were compared: (1) a single-modal student trained from scratch with CE loss as the baseline (Scratch-Student); (2) a student with cross-modal KD from a regular multimodal teacher (Regular-Student), which does not include modality-common encoder and decoders for disentanglement; and (3) a student with cross-modal KD from MDT (the proposed MDT-Student), which performs representation disentanglement and focuses on modality-common information in the KD process. The three student models shared an identical network architecture, comprising a feature extractor, an encoder, and a classification layer, all of which follow those of the teacher model. The hyperparameters were also kept the same for a fair comparison.

For model training, we generated *N*_*com*_ = 1000 complete modality samples with (**X**^*a*^, **X**^*b*^, **y**) and *N*_*incom*_ = 1000 incomplete modality samples featuring only (**X**^*b*^, **y**). The student models were trained on the concatenated (**X**^*b*^, **y**) samples from both complete and incomplete modality datasets; and their respective teachers were trained on complete modality datasets. For model validation and testing, we generated datasets with 1000 samples of (**X**^*a*^, **X**^*b*^, **y**). Evaluation of the student models was based on (**X**^*b*^, **y**) alone. All experiments were conducted with 30 different random replications and the average scores of area under the receiver operating characteristics curve (AUROC) of the test set were reported.

Results of the student models are shown in Table I. MDT-Student and Regular-Student outperformed the student trained from scratch, demonstrating that teacher-to-student KD can improve the predictive performance of the student. The MDT-Student significantly outperformed the Regular-Student. This improvement increased as the amount of overlapping information between the two modalities increased (i.e., *γ* = 0.73).

**TABLE I:** Test AUROC scores comparing MDT-Student with two other student models under varying proportions of modality-common representation (*γ*).

| $\gamma$ | Scratch-Student | Regular-Student | MDT-Student |
| --- | --- | --- | --- |
| 0.07 | 0.7817 | 0.7790 | **0.7957** |
| 0.20 | 0.7898 | 0.7944 | **0.8140** |
| 0.33 | 0.8019 | 0.8039 | **0.8263** |
| 0.47 | 0.8093 | 0.8134 | **0.8318** |
| 0.60 | 0.8121 | 0.8144 | **0.8366** |
| 0.73 | 0.8147 | 0.8180 | **0.8425** |

### D. Teacher Performance Evaluation

The reverse KD is intended to improve the teacher’s performance by learning from the feature extraction capability of the student. This can be beneficial when the teacher has a small training size. To validate this, we evaluated the performance of the MKD teacher (MDT^+^) with a regular teacher (Regular) under varying incompleteness levels. MDT^+^ is initialized with pre-trained weights from the student model’s feature extractor of 𝒳^*b*^. The architecture of the student remains the same as Section III-B. For a fair comparison, all teacher models were coupled with a student model trained from scratch to eliminate the impact of the forward KD process. We set *N*_*com*_ to 500 and varied *N*_*incom*_, exploring values of 250, 500, and 1000 to compare the models’ capability in handling incomplete multimodal data.

Table II compares the performance of the two teacher models. The MDT^+^ displayed a consistent increasing trend in performance when either *N*_*incom*_ or *γ* increases. When *N*_*incom*_ is at its highest, the benefit of reverse KD in MDT^+^ becomes especially pronounced. For smaller values of *N*_*incom*_, the role of *γ* was more critical. A higher *γ* yielded a pronounced advantage when employing the reverse KD in MDT^+^. These and previous results suggest that the effectiveness of both forward and reverse KD in the MKD framework improve with an increased amount of modality-common information. Thus, effectively extracting this overlapping information from different modalities is essential.

**TABLE II:** Test AUROC of MDT^+^ and a regular teacher across varying *N*_*incom*_ and *γ*. Numbers in parentheses denote the corresponding *N*_*incom*_. For each *γ*, the highest AUROC is in **bold**. The values of MDT^+^ that win over those of the regular teacher are underlined.

| $\gamma$ | Regular | MDT<sup>+</sup> (250) | MDT<sup>+</sup> (500) | MDT<sup>+</sup> (1000) |
| --- | --- | --- | --- | --- |
| 0.07 | 0.8192 | 0.7733 | 0.7740 | **0.8369** |
| 0.20 | 0.8221 | 0.8167 | 0.8182 | **0.8427** |
| 0.33 | 0.8339 | 0.8242 | 0.8260 | **0.8439** |
| 0.47 | 0.8336 | <u>0.8349</u> | <u>0.8377</u> | **0.8451** |
| 0.60 | 0.8358 | <u>0.8382</u> | <u>0.8430</u> | **0.8476** |
| 0.73 | 0.8332 | <u>0.8430</u> | <u>0.8458</u> | **0.8513** |

Next, we visualized the representations obtained from MDT^+^ to verify the model’s structural characteristics. Figure 4 presents the results of visualizing the latent representations obtained from the test set using Uniform Manifold Approximation and Projection (UMAP) [38]. We observe that modality-common representations derived from different modalities were located in nearby spaces, whereas modality-specific representations were distinctly separated from the others, confirming disentanglement of modality-common and -specific representations.

**Fig. 4:**
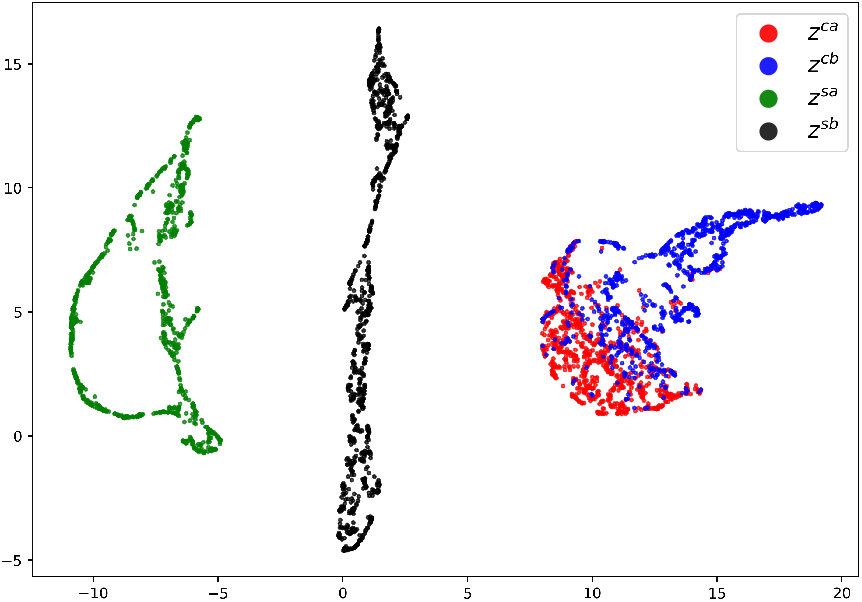
UMAP visualization of the modality-common (**z**^*ca*^, **z**^*cb*^) and modality-specific (**z**^*sa*^, **z**^*sb*^) representations in the simulation test set.

## V. Application IN Early Detection of AD

### A. Data Description and Preprocessing

#### Introduction to ADNI

Data used in the preparation of this article were obtained from the Alzheimer’s Disease Neuroimaging Initiative (ADNI) database (adni.loni.usc.edu). The ADNI was launched in 2003 as a public-private partnership, led by Principal Investigator Michael W. Weiner, MD. The primary goal of ADNI has been to test whether serial MRI, PET, other biological markers, and clinical and neuropsychological assessment can be combined to measure the progression of MCI and early AD. For up-to-date information, see www.adni-info.org.

#### Data Preprocessing

We downloaded 1,310 T1-weighted MRI and 614 FBP-PET images from 961 MCI patients. FBPPET is a type of PET used in AD research for analyzing amyloid plaques in brains, and it can also contribute to predicting the progression from MCI to AD. The MRI scans were spatially normalized by the Computational Anatomy Toolbox 12 (CAT12) [39] with Statistical Parametric Mapping (SPM12) [40] and the Montreal Neuroimaging Institute (MNI) brain atlas. Each PET scan was co-registered to the corresponding MRI. The normalized scans have a size of 121 *×* 121 *×* 121 with a voxel size of 1.5mm in height, width, and depth. The scans were processed with zero padding and resizing to finally achieve a size of 72 *×* 72 *×* 72. The intensity of each scan has been scaled to range from 0 to 1 to facilitate and stabilize model training [14], [41]. The complete dataset consists of 614 pairs of MRI and PET images (*N*_*com*_), and the incomplete dataset consists of 696 MRI-only images (*N*_*incom*_). The patients who converted to AD within 36 months were assigned as converters, otherwise as non-converters. The overall conversion rate in our dataset was 36.6% (480 out of 1,310 scans). We randomly split the paired images into 80% for training, 10% for validation, and 10% for testing while preserving the class distribution. We also ensured that the images from the same patient did not appear in multiple sets, thus preventing data leakage and maintaining the integrity of our evaluation process.

### B. Model Architecture and Training Hyperparameters

The architecture of the MDT is comprised of several distinct components. The feature extractor leverages a ResNet-50 backbone, followed by a single-layer module characterized by a 128-dimensional output, Leaky ReLU activation, and subsequent Layer Normalization. The encoder is designed with a sequence of a 64-dimensional layer, Layer Normalization, a sigmoid activation, followed by another 64-dimensional layer. The decoder consists of a 64-dimensional layer employing a sigmoid activation, followed by Layer Normalization, and concludes with a 128-dimensional layer. We employed a straightforward single-layer classifier.

The training hyperparameters were determined through a grid search on the validation set, ensuring optimal performance. Across all models, the AdamW optimizer was employed with a weight decay of 0.0001 and a half-cosine learning rate scheduling. The batch size was set to 16. The MDT was trained for 100 epochs with a learning rate of 0.001, while the MDT-Student was trained for 30 epochs with a more conservative learning rate set at 0.0001. The MDT^+^ adhered to the same hyperparameters as training the MDT. Following the same hyperparameter tuning procedure in Appendix C, we set *α*_*sim*_ = 10.0, *α*_*diff*_ = 5.0, *α*_*recon*_ = 10.0, *α*_*s*_ = 100.0, and *α*_*t*_ = 25.0. *τ* was set to 5.0.

For data augmentation, we employed random flipping and random rotation. Due to the inherent computational challenges associated with 3D images, we pivoted to a strategy of generating 2D slices [42]. Slices were derived from sagittal, coronal, and axial orientations. Starting from the central point of each orientation, we evenly extracted slices at three-voxel intervals, obtaining 11 slices from each orientation. A total of 33 slices were extracted from each 3D image. All operations during the training phase were performed on these 2D slices. During the test phase, model performance was aggregated as the average value of the logit of each 3D image for patient-level evaluation.

To evaluate the robustness and effectiveness of MKD in our proposed framework, we conducted experiments under various data incomplete ratios (*N*_*incom*_*/*(*N*_*incom*_ + *N*_*com*_)). Although the original training dataset had an incomplete ratio of 0.33, we intentionally increased the incomplete ratio to 0.50 and 0.70 to evaluate the models under settings where the number of PET images is substantially fewer compared to MRIs. This adjustment was achieved by arbitrarily removing PET images from the training set.

### C. Competing Methods

The teacher model is used for classification under a multi-modal setting, whereas the student model can be employed for test patients with only MRI available. Thus, it is important to evaluate both the teacher and the student to understand model’s predictive performance for different patient sub-cohorts.

#### Teacher Comparison

We compared our teacher model (MDT^+^) with various state-of-the-art multimodal learning methods. The competing methods are primarily categorized based on their use of cross-modal KD and their ability to handle incomplete multimodal data. The competing methods were as follows:

- Multi-Late: a complete multimodal model that merges the decisions derived from individual modality models [43];
- Multi-Interm: a complete multimodal model that merges marginal representations from different modalities [43];
- Stagewise: a complete multimodal fusion model that sequentially fuses representations extracted from each modality [44];
- Multitask: a multitask learning model for incomplete multimodal data [45];
- Dual-KD: an incomplete cross-modal KD method that trains two separate teachers on each modality and transfers knowledge to a multimodal student [17];
- Disentangle-KD: an incomplete cross-modal KD framework that imputes missing representation via disentanglement [14].

#### Student Comparison

The student model is intended for the use case of predicting for patients with only MRI scans. There are a variety of choices to train such a model. A straightforward approach is to train a single-modal model using only MRI. Another option is to use multimodal data in training and single-modal data at test time.

First, we compared the MDT-Student with the following methods on the original dataset:

- ResNet-MRI: a conventional ResNet-50 model trained with only MRI;
- Patch-MRI: a cross-modal KD model with complete multimodal dataset that utilizes multiple patches of a whole brain to capture region-specific features [21];
- Stagewise-MRI: the MRI component of this complete multimodal framework was used [44].

These methods either exclude samples with missing modalities or exclude modalities with missing samples to train the student. The purpose of this comparison is to show the benefit of incomplete multimodal learning even when the test patient does not possess the modality with incomplete data (PET).

Then, we compared MDT-Student with the following incomplete multimodal methods at varied incomplete ratios:

- S2S-KD: single-to-single cross-modal KD from PET to MRI;
- M2S-KD: multi-to-single cross-modal KD from MRI & PET to MRI;
- Multitask-MRI [45];
- Dual-KD-MRI [17];
- Disentangle-KD-MRI [14].

For S2S-KD and M2S-KD, we used KL-divergence loss only for the complete dataset and CE loss for the incomplete dataset. For other methods, we used their respective MRI components. The hyperparameters of all competing methods were tuned via grid search, starting with the suggested ranges in the original papers and extending beyond when necessary to ensure fair comparison.

### D. Experimental Results

#### Student Evaluation

We start by comparing the MDT-Student on the dataset with the original incomplete ratio with competing methods that cannot handle incomplete modality data (Table III). The single-modal model, ResNet-MRI, underperformed all other multimodal models. This highlights the valuable role of PET in enhancing AD diagnosis accuracy and the benefit of multimodal learning. The MDT-Student significantly outperformed the complete multimodal methods, with an AUROC that was 0.058 higher than Patch-MRI. This notable performance enhancement stems from the model’s capability of leveraging incomplete multimodal data.

**TABLE III:** Classification performance with only MRI. The competing methods include models trained exclusively with MRI (ResNet-MRI) and complete multimodal models (Stagewise-MRI and Patch-MRI). The best model is in **bold**.

| Model | AUROC | Accuracy | Sensitivity | Specificity |
| --- | --- | --- | --- | --- |
| ResNet-MRI | 0.7110 | 0.6999 | 0.6457 | 0.7183 |
| Stagewise-MRI | 0.7215 | 0.7025 | 0.6483 | 0.7196 |
| Patch-MRI | 0.7376 | 0.7216 | 0.6537 | 0.7336 |
| MDT-Student | **0.7956** | **0.7639** | **0.6803** | **0.7864** |

Then, we compared the classification performance of the MDT-Student model with incomplete multimodal methods at varied incomplete ratio levels (Table IV). Overall, the MDT-Student consistently outperformed all others across all levels of incomplete ratio, demonstrating the benefit of representation disentanglement in cross-modal KD. Among the competing methods, S2S-KD consistently underperformed, likely due to its reliance solely on PET data for the teacher model, which limits its ability to leverage complementary information from MRI. M2S-KD showed competitive performance at lower incomplete ratios but struggled as the amount of available PET data decreased, highlighting the challenge of effectively distilling knowledge when multimodal data is scarce.

**TABLE IV:**
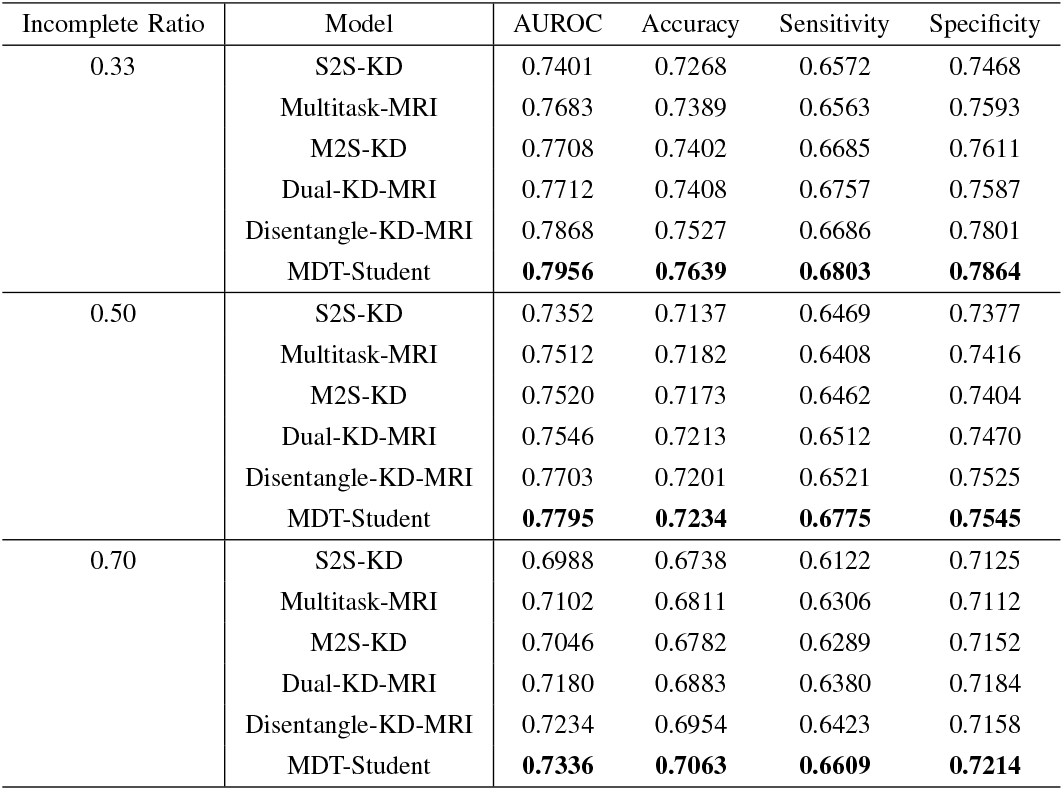
Classification performance with only MRI under various incomplete ratios. The competing methods are incomplete multimodal models. The best model in each incomplete ratio is in **bold**.

Notably, at an incomplete ratio of 0.70 where the amount of PET data was significantly reduced, the performance gap between MDT-Student and the competing methods widened. This underscores that effective KD of modality-common information can significantly mitigate performance degradation even at high missing rates.

Moreover, the performance of the MDT-Student at a high incomplete ratio still remained on par with models run in the complete multimodal setting such as Stagewise-MRI and Patch-MRI (Table III). In particular, MDT-Student achieved an AUROC of 0.7336 when 70% of training patients missed PET, while Stagewise-MRI and Patch-MRI achieved an AUROC of 0.7215 and 0.7376, respectively. These results highlight the benefits of the proposed method in AD diagnosis when PET data is available for a small portion of patients.

Last but not least, Disentangle-KD, like our MDT-Student, employs representation disentanglement and cross-modal KD. However, it differs in its testing phase, using one model for both MRI-only and combined MRI-PET cases. Our framework produces two models, one tailored for each use case, leading to improved performance over Disentangle-KD. This demonstrates the effectiveness of specialized models over onesize-fits-all models in diverse modality availability scenarios. These results highlight the potential of our method to improve healthcare accessibility, especially for patients without access to advanced diagnostic imaging like PET scans. By effectively leveraging knowledge from limited PET data and applying it to more widely available MRI data, our approach could enable more accurate AD diagnosis for a broader patient population.

#### Teacher Evaluation

Table V presents the results of teacher models tested on patients with both MRI and PET data. Although different incomplete ratios were tested, the complete multimodal methods, Multi-Late, Multi-Interm, and Stagewise, were trained exclusively using complete data from MRI and PET scans. To further investigate the discriminative power of individual modalities, we also conducted experiments using a PET-only model (ResNet-PET). ResNet-PET achieved an AUROC of 0.8272, accuracy of 0.7813, sensitivity of 0.7264, and specificity of 0.7986. This performance falls between that of MRI-only and multimodal models, confirming the high discriminative capability of PET for AD conversion.

**TABLE V:** Classification performance with MRI and PET under various incomplete ratios. Models marked with * indicate incomplete multimodal models, while others represent complete multimodal models. The best model in each incomplete ratio is in **bold**.

| Incomplete Ratio | Model | AUROC | Accuracy | Sensitivity | Specificity |
| --- | --- | --- | --- | --- | --- |
| 0.33 | Multi-Late | 0.8623 | 0.8257 | 0.7709 | 0.8126 |
|  | Multi-Interm | 0.8693 | 0.8285 | 0.7763 | 0.8387 |
|  | Stagewise | 0.8699 | 0.8290 | 0.7790 | 0.8402 |
|  | Multitask* | 0.8720 | 0.8325 | 0.7822 | 0.8406 |
|  | Dual-KD* | 0.8751 | 0.8378 | 0.7863 | 0.8433 |
|  | Disentangle-KD* | 0.8783 | 0.8389 | 0.7890 | 0.8467 |
|  | MDT<sup>+</sup>* | **0.8815** | **0.8428** | **0.7918** | **0.8484** |
| 0.50 | Multi-Late | 0.8507 | 0.8142 | 0.7619 | 0.8368 |
|  | Multi-Interm | 0.8534 | 0.8187 | 0.7666 | 0.8372 |
|  | Stagewise | 0.8549 | 0.8188 | 0.7637 | 0.8378 |
|  | Multitask* | 0.8652 | 0.8225 | 0.7721 | 0.8397 |
|  | Dual-KD* | 0.8653 | 0.8250 | 0.7730 | 0.8403 |
|  | Disentangle-KD* | 0.8690 | 0.8275 | 0.7856 | 0.8438 |
|  | MDT<sup>+</sup>* | **0.8782** | **0.8370** | **0.7927** | **0.8531** |
| 0.70 | Multi-Late | 0.8301 | 0.7887 | 0.7424 | 0.8188 |
|  | Multi-Interm | 0.8342 | 0.7927 | 0.7450 | 0.8164 |
|  | Stagewise | 0.8397 | 0.8002 | 0.7498 | 0.8204 |
|  | Multitask* | 0.8338 | 0.7949 | 0.7461 | 0.8109 |
|  | Dual-KD* | 0.8473 | 0.8055 | 0.7531 | 0.8167 |
|  | Disentangle-KD* | 0.8511 | 0.8079 | 0.7535 | 0.8267 |
|  | MDT<sup>+</sup>* | **0.8663** | **0.8201** | **0.7627** | **0.8365** |

Across all incomplete ratios, the MDT^+^ consistently outperformed competing methods. The Multi-Late model had the lowest performance. This suggests that models capable of considering interrelationships between modalities are more effective than ensembling models trained separately on each modality. Furthermore, incomplete multimodal methods, Multitask, Dual-KD, Disentangle-KD, and MDT^+^, achieved higher performances than complete multimodal methods, demonstrating the benefit of using the additional MRI samples.

The performance advantage of MDT^+^ is particularly notable at higher incomplete ratios, where reverse KD plays a crucial role. Although the teacher model already utilizes MRI data, reverse KD provides complementary MRI knowledge from a larger MRI dataset. This is especially beneficial when only limited complete samples are available, i.e., at high incomplete ratios. Similar to the previous MRI-only experiment, we observed less performance degradation of our proposed method at increased incomplete ratios compared to other models. The highest performance gap was observed at an incomplete ratio of 0.70, aligning with the MRI-only results in Table IV. This suggests that our proposed framework is particularly beneficial for AD applications with a high modality missing rate. Note that the improvement gap was less pronounced compared to the MRI-only experiment as PET carries more critical diagnosis information than MRI (Table IV).

The evaluation results of teacher and student models show that the MKD process benefited both the MRI-only model and the multimodal MRI&PET model, achieving improved performance for both prediction tasks. It is worth noting that the improvement gap was less pronounced compared to the MRI-only experiment, as PET carries more critical diagnostic information than MRI (Table IV). However, the consistent outperformance of MDT^+^ across all incomplete ratios underscores the value of our mutual KD approach in incomplete multimodal settings, even when one modality (PET) is particularly informative.

## VI. Conclusions

In this paper, we addressed the challenge of early detection of AD by developing a framework utilizing MKD for predictive modeling across patient sub-cohorts with varying imaging modalities. The bi-directional nature of our method facilitates a mutual exchange of knowledge between the student and the teacher. The MDT model, the core of our approach, strategically focuses on modality-common information for efficient KD. We further enhanced the teacher by distilling the feature extraction knowledge of the student. Our theoretical analysis reinforces the benefits of representation disentanglement in multi-to-single cross-modal KD. These theoretical insights were supported by simulation studies. Our model significantly outperformed the student of a regular multimodal teacher, especially with a higher proportion of modality-common information. Applied to a case study in the early AD diagnosis, our method showed substantial performance gains for patients with both multimodal and single-modal data across varying incompleteness levels.

The practical impact of our method on healthcare is significant. While PET imaging is valuable for AD diagnosis, its use is often limited by cost and insurance issues. Our method could benefit patients unable to access PET scans, enabling early AD diagnosis and better treatment planning for a broader population. By making advanced AD diagnostic capabilities more accessible, we potentially improve outcomes for many patients who might otherwise miss out on early intervention opportunities. This aligns with the growing need for cost-effective, widely available diagnostic tools in AD management.

While our model showed promising results, there are several limitations and intriguing directions to extend our work. First, even if we provide an efficient tuning strategy, there are many hyperparameters that require time-intensive tuning to find their optimal values. This indicates the need for simplification, possibly by streamlining the model structure or combining the similarity and difference loss components. Another direction is extending MKD to settings with more than two modalities. We could apply our method to modality pairs, creating up to *M* (*M* − 1)*/*2 pair-wise MDT models for *M* modalities. These models’ outputs could be combined via ensemble learning techniques. Student models could then be trained for individual modalities, learning from the ensemble of teachers. This approach would use our current method as a building block for multiple modalities, preserving bidirectional KD and modality disentanglement benefits while scaling to more complex scenarios. Furthermore, the concept of extracting modality-common representations can be developed into a strategy to mitigate distribution shifts across different modalities. This would involve projecting features into a unified modality-common representation space, thereby reducing discrepancies and enhancing the model’s adaptability and performance across varied modalities. Last but not least, developing interpretability methods for the model’s decisions is crucial for clinical adoption and trust. This involves techniques to discover the key brain regions contributing to predictions, helping clinicians understand and validate the model’s reasoning.

## Data Availability

All data produced are available online at the Alzheimer's Disease Neuroimaging Initiative (ADNI) database.

https://adni.loni.usc.edu/

## Acknowledgments

This research was funded by NIH grant 2R42AG053149-02A1 and NSF grant DMS-2053170. This research was also supported by NIH grants R01AG069453 and P30AG072980, the State of Arizona, and Banner Alzheimer’s Foundation. Data collection and sharing for this project was funded by the Alzheimer’s Disease Neuroimaging Initiative (ADNI) (National Institutes of Health Grant U01 AG024904) and DOD ADNI (Department of Defense award number W81XWH-12-2-0012). ADNI is funded by the National Institute on Aging, the National Institute of Biomedical Imaging and Bioengineering, and through generous contributions from the following: AbbVie, Alzheimer’s Association; Alzheimer’s Drug Discovery Foundation; Araclon Biotech; BioClinica, Inc.; Biogen; Bristol-Myers Squibb Company; CereSpir, Inc.; Cogstate; Eisai Inc.; Elan Pharmaceuticals, Inc.; Eli Lilly and Company; EuroImmun; F. Hoffmann-La Roche Ltd and its affiliated company Genentech, Inc.; Fujirebio; GE Healthcare; IXICO Ltd.; Janssen Alzheimer Immunotherapy Research & Development, LLC.; Johnson & Johnson Pharmaceutical Research & Development LLC.; Lumosity; Lundbeck; Merck & Co., Inc.; Meso Scale Diagnostics, LLC.; NeuroRx Research; Neurotrack Technologies; Novartis Pharmaceuticals Corporation; Pfizer Inc.; Piramal Imaging; Servier; Takeda Pharmaceutical Company; and Transition Therapeutics. The Canadian Institutes of Health Research is providing funds to support ADNI clinical sites in Canada. Private sector contributions are facilitated by the Foundation for the National Institutes of Health (www.fnih.org). The grantee organization is the Northern California Institute for Research and Education, and the study is coordinated by the Alzheimer’s Therapeutic Research Institute at the University of Southern California. ADNI data are disseminated by the Laboratory for Neuro Imaging at the University of Southern California.

## Appendix A

### Proof OF Theorem 1

It would suffice to prove that

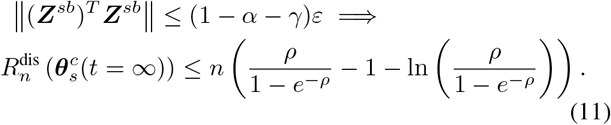

First, we prove that there exists an 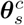 such that 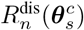 is bounded. Suppose a trained teacher with parameter 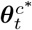 The least squares estimator (LSE) that minimizes the squared difference between the student and teacher is:

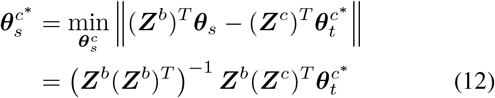

Without loss of generality, assume 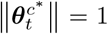 as scaling the coefficients does not affect predictions in a linear model. Then,

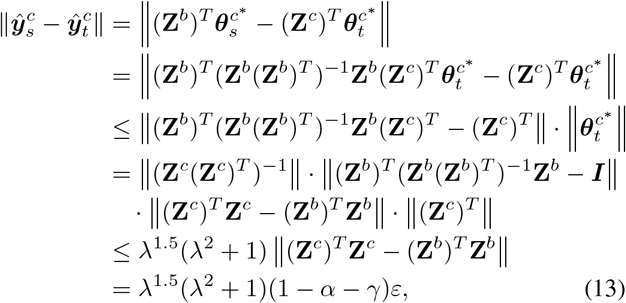

where the second last line comes from

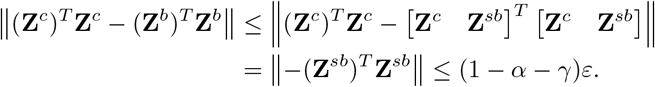

Applying Lemma 1 and Corollary 1 from [34] to (13) and the definition of KD loss, we obtain the following upper bound for the empirical KD loss:

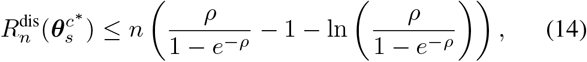

where *ρ* = *λ*^1.5^(*λ*^2^ + 1)(1 − *α* − *γ*). The right-hand side is a monotonically increasing function when *ρ* is positive. Next, we prove that if the training process is long enough, *t* → ∞, then the final KD loss is upper bounded by 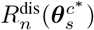.

Since 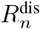 is convex in the pair of probability distributions, the gradient drives the objective value closer to the optimal minimum in each iteration. Namely, the objective value monotonically decreases along *t*. Thus, for any *t* ∈ [0, ∞), the gradient of 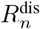 is negative,

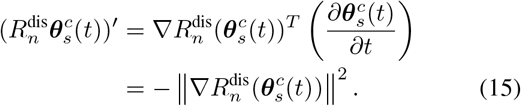

We apply a modified version of restricted Polyak-Lojasiewicz inequality from Corollary A.1 in [46] with a slight modification of replacing the minimum value of 0 with 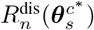. For any sublevel set 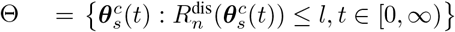 such that

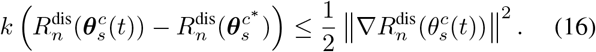

This inequality lower bounds the reduction in the objective in terms of the gap between the objective and the optimum, which proves convergence. Flipping the inequality leads to

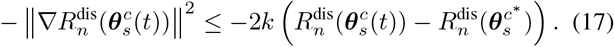

Combining with (14), we have an upper bound for the gradient of 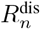 for *k >* 0 and *t* ∈ [0, ∞) as follows:

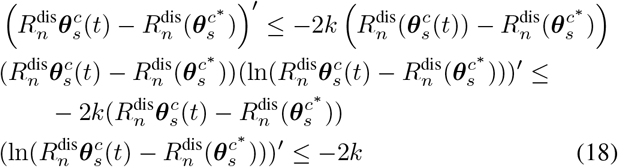

Integrating (18) over [0, *t*),

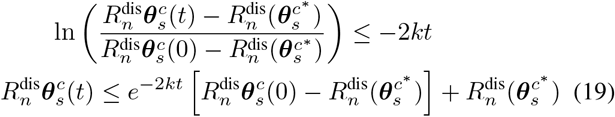

As *t* → *∞, e*^*™2kt* → *0*, we have^

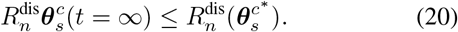

Thus, we showed that if the training process is long enough, then the empirical KD loss is upper bounded by 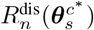, a monotonic function with respect to 1 − *α* − *γ*. Given *α*, as *γ* increases, the upper bound of distillation loss decreases. ◼

## Appendix B

### Proof OF Theorem 2

The predictive equations for the regular teacher and its student are 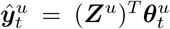 and 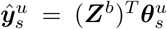. Suppose the regular teacher has trained parameter 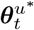. Then, the LSE of the corresponding student is as follows:

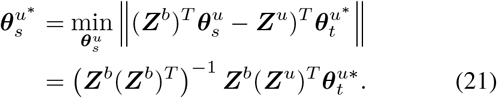

Following the same algebra as in the proof of Theorem 1, we obtain an upper bound for the mean squared error between the student and the regular teacher’s predictions as follows:

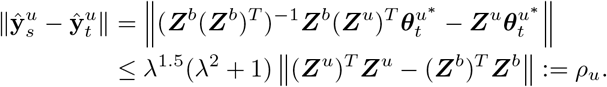

Since Z^u^ = [Z^c^ Z^sa^ Z^sb^], it is clear that

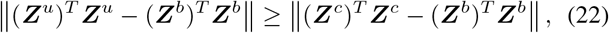

which leads to *ρ*^*u*^ ≥ *ρ*^*c*^.

Consequently, following the same steps in the proof of Theorem 1, we have

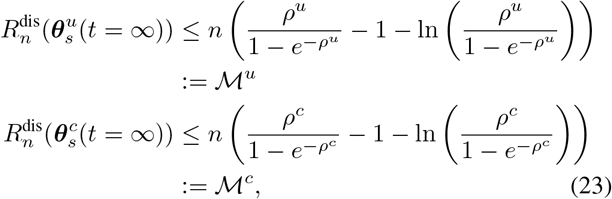

and ℳ^*u*^ ≥ ℳ^*c*^. This implies that the student model trained with MDT has a tighter upper bound on empirical KD loss than the student model trained with regular teacher. ◼

## Appendix C

### Hyperparameter Search IN Simulation Study

We introduce an efficient tuning strategy focused on the most crucial aspect of the MDT, which is effective disentanglement. We conducted a grid search to tune the balancing hyperparameters of the MKD framework. For simplicity, we fixed *d*_*c*_ to 10 and *N*_*incom*_ to 1000. The hyperparameters *α*_*sim*_ and *α*_*diff*_ directly impact disentanglement. As shown in Figure 5a, setting these values too high or too low decreases model performance. We selected *α*_*sim*_ = 2.0 and *α*_*diff*_ = 1.0, which yielded the best performance.

**Fig. 5:**
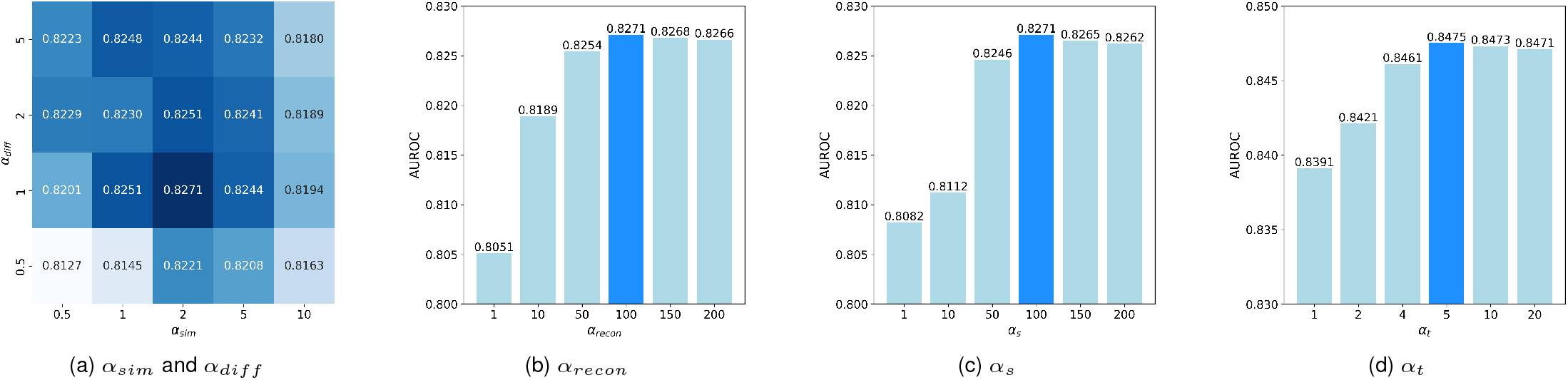
Hyperparameter tuning results for MKD. (a) *α*_*sim*_ and *α*_*diff*_ for MDT-Student, (b) *α*_*recon*_ for MDT-Student, (c) *α*_*s*_ for MDT-Student, and (d) *α*_*t*_ for MDT^+^. The highest AUROC value for each experiment is highlighted in dark blue.

For the remaining hyperparameters (*α*_*recon*_, *α*_*s*_, and *α*_*t*_), we observed the importance of aligning their scales with the CE loss. The CE losses for both MDT-Student and MDT^+^ models were approximately 0.1, while *ℒ* _*recon*_, *ℒ* _*KL*_, and 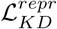 were 0.0007, 0.0015, and 0.027, respectively. Figures 5b, 5c, and 5d demonstrate that multiplying *α*_*recon*_, *α*_*s*_, and *α*_*t*_ by their respective loss terms to match the CE loss scale stabilizes performance. Values that are too small may result in reconstruction regularization or KD effects being too weak compared to the classification task, potentially compromising the intended benefits. We selected *α*_*recon*_ = 100, *α*_*s*_ = 100, and *α*_*t*_ = 5 based on these observations.

Moreover, we conducted experiments to verify the representation regularization role of the decoder. We maintained the obtained hyperparameter values, except *α*_*recon*_ = 0.0 for comparison. To assess the model’s training progression, we monitored the validation CE loss. As illustrated in Figure 6, the CE loss for the MDT model with a decoder exhibits a steady decline and convergence. However, the MDT model without a decoder begins to exhibit overfitting, with the CE loss increasing after 30 epochs. Such patterns can serve as evidence of degenerated representations, highlighting the importance of representation regularization.

**Fig. 6:**
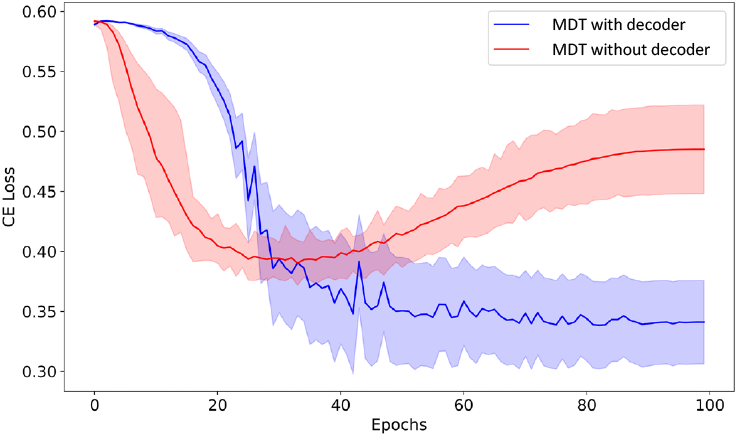
Comparisons of validation CE loss for MDT models with (blue) and without (red) decoder. Shaded regions represent standard deviations above and below the respective mean values. MDT without decoder overfits, with the CE loss increasing after 30 epochs.

---

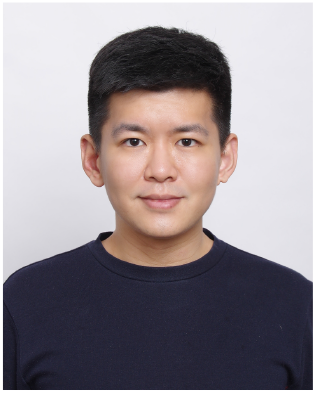

**Min Gu Kwak** received his B.S. and Ph.D. degrees in the Department of Industrial and Management Engineering at Korea University, Seoul, Republic of Korea, in 2016 and 2022, respectively. He is currently a Postdoctoral Fellow in the H. Milton Stewart School of Industrial and Systems Engineering at Georgia Institute of Technology, GA, USA. His research primarily focuses on using deep learning methods for out-of-distribution or limited labeled data. The application domains mainly include multichannel sensors, images, manufacturing processes, and medical data. He is a member of INFORMS.

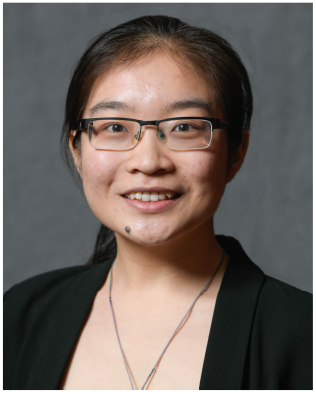

**Lingchao Mao** received her B.S. degrees in Statistics and Industrial & Systems Engineering in 2020 from North Carolina State University. She is now working towards her Ph.D. degree in Machine Learning at Georgia Institute of Technology advised by Dr. Jing Li. Her research interests include statistical methods and machine learning for healthcare applications. She is a member of INFORMS and IISE.

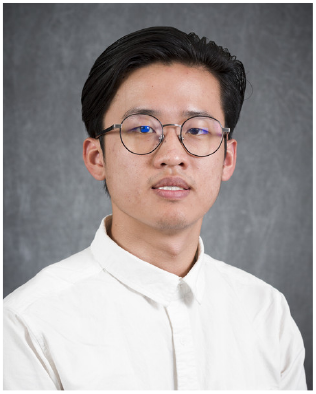

**Zhiyang Zheng** received the B.S. degree in statistics from the University of Science and Technology of China, Hefei, China, in 2018. He is currently pursuing the Ph.D. degree with the H. Milton Stewart School of Industrial and Systems Engineering, Georgia Institute of Technology, Atlanta, GA, USA. He is currently a Research Assistant with the Georgia Institute of Technology, supervised by Dr. Jing Li. His research interest includes medical image-based machine learning and deep learning.

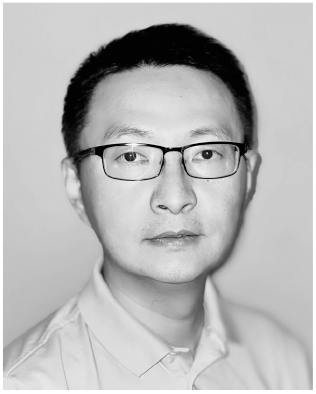

**Yi Su** received his Ph.D. degree in Biomedical Engineering from Mayo Graduate School at Rochester, MN. He leads the Computational Image Analysis Laboratory at the Banner Alzheimer’s Institute in Phoenix, AZ. He also directs/co-directs the Data Management and Statistics and Biomarker Cores at the Arizona Alzheimer’s Disease Research Center. His research focuses on the development and application of novel imaging and data analytics techniques to better understand aging and neurodegenerative diseases such as Alzheimer’s disease.

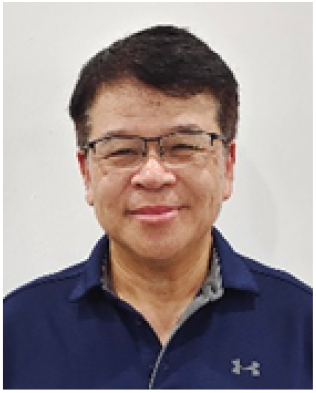

**Fleming Lure** received his Ph.D. degree in Electrical Engineering from the Pennsylvania State University, State College, PA. He is a Chief Product Officer in MS Technologies Corp, Rockville, MD, USA. His research interests are computer aided detection and machine learning for disease detection, diagnosis, and prognosis. He has led a team to receive the first FDA-approved early-stage lung cancer detection system on radiograph. He is a member of Radiological Society of North America (RSNA).

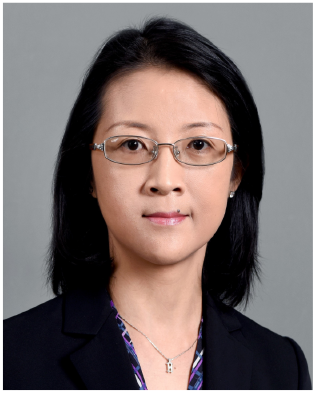

**Jing Li** received her Ph.D. degree in Industrial and Operations Engineering from the University of Michigan at Ann Arbor, MI. She is a Professor in the School of Industrial and Systems Engineering, Georgia Institute of Technology, Atlanta, GA, USA. Her research interests are statistical modeling and machine learning for health care applications. She is a recipient of NSF CAREER award. She is a member of IISE, INFORMS, and IEEE.

## References

[1] S. Roy, J. Wang, and Y. Xu, “Alzheimer’s disease facts and figures,” Alzheimers Dement, vol. 19, pp. 1598–1695, 2023.

[2] V. A. Canady, “Fda approves new treatment for alzheimer’s disease,” Mental Health Weekly, vol. 33, no. 3, pp. 6–7, 2023.

[3] J. R. Sims, J. A. Zimmer, C. D. Evans, M. Lu, P. Ardayfio, J. Sparks, A. M. Wessels, S. Shcherbinin, H. Wang, E. S. M. Nery et al., “Donanemab in early symptomatic alzheimer disease: the trailblazer-alz 2 randomized clinical trial,” Jama, vol. 330, no. 6, pp. 512–527, 2023.

[4] K. G. Yiannopoulou and S. G. Papageorgiou, “Current and future treatments in alzheimer disease: an update,” Journal of central nervous system disease, vol. 12, p. 1179573520907397, 2020.

[5] K.-H. Thung, C.-Y. Wee, P.-T. Yap, and D. Shen, “Identification of progressive mild cognitive impairment patients using incomplete longitudinal mri scans,” Brain Structure and Function, vol. 221, pp. 3979– 3995, 2016.

[6] D. P. Veitch, M. W. Weiner, P. S. Aisen, L. A. Beckett, C. DeCarli, R. C. Green, D. Harvey, C. R. Jack Jr, W. Jagust, S. M. Landau et al., “Using the alzheimer’s disease neuroimaging initiative to improve early detection, diagnosis, and treatment of alzheimer’s disease,” Alzheimer’s & Dementia, vol. 18, no. 4, pp. 824–857, 2022.

[7] X. Liu, K. Chen, T. Wu, D. Weidman, F. Lure, and J. Li, “Use of multi-modality imaging and artificial intelligence for diagnosis and prognosis of early stages of alzheimer’s disease,” Translational Research, vol. 194, pp. 56–67, 2018.

[8] Y. Pan, M. Liu, C. Lian, T. Zhou, Y. Xia, and D. Shen, “Synthesizing missing pet from mri with cycle-consistent generative adversarial networks for alzheimer’s disease diagnosis,” in Medical Image Computing and Computer Assisted Intervention–MICCAI 2018: 21st International Conference, Granada, Spain, September 16-20, 2018, Proceedings, Part III 11. Springer, 2018, pp. 455–463.

[9] E. K. Degenhardt, M. M. Witte, M. G. Case, P. Yu, D. B. Henley, H. M. Hochstetler, D. N. D’Souza, and P. T. Trzepacz, “Florbetapir f18 pet amyloid neuroimaging and characteristics in patients with mild and moderate alzheimer dementia,” Psychosomatics, vol. 57, no. 2, pp. 208–216, 2016.

[10] Y. Liu, L. Fan, C. Zhang, T. Zhou, Z. Xiao, L. Geng, and D. Shen, “Incomplete multi-modal representation learning for alzheimer’s disease diagnosis,” Medical Image Analysis, vol. 69, p. 101953, 2021.

[11] H. Ye, Q. Zhu, Y. Yao, Y. Jin, and D. Zhang, “Pairwise feature-based generative adversarial network for incomplete multi-modal alzheimer’s disease diagnosis,” The Visual Computer, vol. 39, no. 6, pp. 2235–2244, 2023.

[12] J. Song, J. Zheng, P. Li, X. Lu, G. Zhu, and P. Shen, “An effective multimodal image fusion method using mri and pet for alzheimer’s disease diagnosis,” Frontiers in digital health, vol. 3, p. 637386, 2021.

[13] F. Zhang, Z. Li, B. Zhang, H. Du, B. Wang, and X. Zhang, “Multi-modal deep learning model for auxiliary diagnosis of alzheimer’s disease,” Neurocomputing, vol. 361, pp. 185–195, 2019.

[14] Y. Chen, Y. Pan, Y. Xia, and Y. Yuan, “Disentangle first, then distill: A unified framework for missing modality imputation and alzheimer’s disease diagnosis,” IEEE Transactions on Medical Imaging, 2023.

[15] X. Gao, F. Shi, D. Shen, and M. Liu, “Multimodal transformer network for incomplete image generation and diagnosis of alzheimer’s disease,” Computerized Medical Imaging and Graphics, vol. 110, p. 102303, 2023.

[16] J. Gou, B. Yu, S. J. Maybank, and D. Tao, “Knowledge distillation: A survey,” International Journal of Computer Vision, vol. 129, no. 6, pp. 1789–1819, 2021.

[17] Q. Wang, L. Zhan, P. Thompson, and J. Zhou, “Multimodal learning with incomplete modalities by knowledge distillation,” in Proceedings of the 26th ACM SIGKDD International Conference on Knowledge Discovery & Data Mining, 2020, pp. 1828–1838.

[18] C. Wang, S. Piao, Z. Huang, Q. Gao, J. Zhang, Y. Li, H. Shan, A. D. N. Initiative et al., “Joint learning framework of cross-modal synthesis and diagnosis for alzheimer’s disease by mining underlying shared modality information,” Medical Image Analysis, vol. 91, p. 103032, 2024.

[19] N. C. Garcia, P. Morerio, and V. Murino, “Modality distillation with multiple stream networks for action recognition,” in Proceedings of the European Conference on Computer Vision (ECCV), 2018, pp. 103–118.

[20] M. Hu, M. Maillard, Y. Zhang, T. Ciceri, G. La Barbera, I. Bloch, and P. Gori, “Knowledge distillation from multi-modal to mono-modal segmentation networks,” in Medical Image Computing and Computer Assisted Intervention–MICCAI 2020: 23rd International Conference, Lima, Peru, October 4–8, 2020, Proceedings, Part I 23. Springer, 2020, pp. 772–781.

[21] H. Guan, C. Wang, and D. Tao, “Mri-based alzheimer’s disease prediction via distilling the knowledge in multi-modal data,” NeuroImage, vol. 244, p. 118586, 2021.

[22] C. Zhang, E. Adeli, T. Zhou, X. Chen, and D. Shen, “Multi-layer multiview classification for alzheimer’s disease diagnosis,” in Proceedings of the AAAI Conference on Artificial Intelligence, vol. 32, no. 1, 2018.

[23] S. Spasov, L. Passamonti, A. Duggento, P. Lio, N. Toschi, A. D. N. Initiative et al., “A parameter-efficient deep learning approach to predict conversion from mild cognitive impairment to alzheimer’s disease,” Neuroimage, vol. 189, pp. 276–287, 2019.

[24] S. Dwivedi, T. Goel, M. Tanveer, R. Murugan, and R. Sharma, “Multimodal fusion-based deep learning network for effective diagnosis of alzheimer’s disease,” IEEE MultiMedia, vol. 29, no. 2, pp. 45–55, 2022.

[25] Y. Shi, H.-I. Suk, Y. Gao, S.-W. Lee, and D. Shen, “Leveraging coupled interaction for multimodal alzheimer’s disease diagnosis,” IEEE transactions on neural networks and learning systems, vol. 31, no. 1, pp. 186–200, 2019.

[26] G. Hinton, O. Vinyals, and J. Dean, “Distilling the knowledge in a neural network,” arXiv preprint 1503.02531, 2015.

[27] Y. Li, J. Luo, and J. Zhang, “Classification of alzheimer’s disease in mri images using knowledge distillation framework: an investigation,” International Journal of Computer Assisted Radiology and Surgery, vol. 17, no. 7, pp. 1235–1243, 2022.

[28] J. Zhu, Y. Tan, R. Lin, J. Miao, X. Fan, Y. Zhu, P. Liang, J. Gong, and H. He, “Efficient self-attention mechanism and structural distilling model for alzheimer’s disease diagnosis,” Computers in Biology and Medicine, vol. 147, p. 105737, 2022.

[29] F. M. Thoker and J. Gall, “Cross-modal knowledge distillation for action recognition,” in 2019 IEEE International Conference on Image Processing (ICIP). IEEE, 2019, pp. 6–10.

[30] M. Rahimpour, J. Bertels, A. Radwan, H. Vandermeulen, S. Sunaert, D. Vandermeulen, F. Maes, K. Goffin, and M. Koole, “Cross-modal distillation to improve mri-based brain tumor segmentation with missing mri sequences,” IEEE Transactions on Biomedical Engineering, vol. 69, no. 7, pp. 2153–2164, 2021.

[31] S. Zhou, W. Liu, C. Hu, S. Zhou, and C. Ma, “Unidistill: A universal cross-modality knowledge distillation framework for 3d object detection in bird’s-eye view,” in Proceedings of the IEEE/CVF conference on computer vision and pattern recognition, 2023, pp. 5116–5125.

[32] M. Ghorbani, M. Bahrami, A. Kazi, M. Soleymani Baghshah, H. R. Rabiee, and N. Navab, “Gkd: Semi-supervised graph knowledge distillation for graph-independent inference,” in Medical Image Computing and Computer Assisted Intervention–MICCAI 2021: 24th International Conference, Strasbourg, France, September 27–October 1, 2021, Proceedings, Part V 24. Springer, 2021, pp. 709–718.

[33] T. Zhou, M. Liu, K.-H. Thung, and D. Shen, “Latent representation learning for alzheimer’s disease diagnosis with incomplete multimodality neuroimaging and genetic data,” IEEE transactions on medical imaging, vol. 38, no. 10, pp. 2411–2422, 2019.

[34] Z. Xue, Z. Gao, S. Ren, and H. Zhao, “The modality focusing hypothesis: Towards understanding crossmodal knowledge distillation,” arXiv preprint 2206.06487, 2022.

[35] K. He, X. Zhang, S. Ren, and J. Sun, “Deep residual learning for image recognition,” in Proceedings of the IEEE conference on computer vision and pattern recognition, 2016, pp. 770–778.

[36] G. Aguilar, Y. Ling, Y. Zhang, B. Yao, X. Fan, and C. Guo, “Knowledge distillation from internal representations,” in Proceedings of the AAAI conference on artificial intelligence, vol. 34, no. 05, 2020, pp. 7350– 7357.

[37] I. Loshchilov and F. Hutter, “Decoupled weight decay regularization,” arXiv preprint 1711.05101, 2017.

[38] L. McInnes, J. Healy, and J. Melville, “Umap: Uniform manifold approximation and projection for dimension reduction,” arXiv preprint 1802.03426, 2018.

[39] C. Gaser, R. Dahnke, P. M. Thompson, F. Kurth, E. Luders, and A. D. N. Initiative, “Cat–a computational anatomy toolbox for the analysis of structural mri data,” biorxiv, pp. 2022–06, 2022.

[40] J. Ashburner, G. Barnes, C.-C. Chen, J. Daunizeau, G. Flandin, K. Friston, S. Kiebel, J. Kilner, V. Litvak, R. Moran et al., “Spm12 manual,” Wellcome Trust Centre for Neuroimaging, London, UK, vol. 2464, no. 4, 2014.

[41] C. Jiang, Y. Pan, Z. Cui, D. Nie, and D. Shen, “Semi-supervised standard-dose pet image generation via region-adaptive normalization and structural consistency constraint,” IEEE transactions on medical imaging, vol. 42, no. 10, pp. 2974–2987, 2023.

[42] M. Liu, D. Cheng, W. Yan, and A. D. N. Initiative, “Classification of alzheimer’s disease by combination of convolutional and recurrent neural networks using fdg-pet images,” Frontiers in neuroinformatics, vol. 12, p. 35, 2018.

[43] S. R. Stahlschmidt, B. Ulfenborg, and J. Synnergren, “Multimodal deep learning for biomedical data fusion: a review,” Briefings in Bioinformatics, vol. 23, no. 2, p. bbab569, 2022.

[44] T. Zhou, K.-H. Thung, X. Zhu, and D. Shen, “Effective feature learning and fusion of multimodality data using stage-wise deep neural network for dementia diagnosis,” Human brain mapping, vol. 40, no. 3, pp. 1001– 1016, 2019.

[45] K.-H. Thung, P.-T. Yap, and D. Shen, “Multi-stage diagnosis of alzheimer’s disease with incomplete multimodal data via multi-task deep learning,” in International Workshop on Deep Learning in Medical Image Analysis. Springer, 2017, pp. 160–168.

[46] M. Phuong and C. Lampert, “Towards understanding knowledge distillation,” in International conference on machine learning. PMLR, 2019, pp. 5142–5151.

